# Impacts of regional lockdown policies on COVID-19 transmission in India in 2020

**DOI:** 10.1101/2021.08.09.21261277

**Authors:** Aarushi Kalra, Paul Novosad

## Abstract

**Objective:** To assess the impact of non-pharmaceutical interventions (NPIs) on the first wave of COVID transmission and fatalities in India.

**Methods:** We collected data on NPIs, using government notifications and news reports, in six major Indian states from March to August 2020, and we matched these with district-level data on COVID related deaths and Google Mobility reports. We used a district fixed effect regression approach to measure the extent to which district-level lockdowns and mobility restrictions helped reduce deaths in 2020.

**Results:** In most states, COVID deaths grew most rapidly only after the initial lockdown was lifted. District-level NPIs were associated with a statistically significantly lower COVID death count in three out of five sample states (district analysis was not possible in Delhi) and in the aggregate. Interventions that were most associated with slowing fatalities were temple closures, retail closures, and curfews.

**Discussion:** Outside of Maharashtra (the first state struck) the first fatality wave appears to have been delayed by the national lockdown. India’s NPIs, however incomplete, were successful in delaying or limiting COVID-19 deaths. Even with incomplete compliance, limiting mass gatherings in face of incipient viral waves may save lives.

## Introduction

The first case of COVID-19 was confirmed in India on January 30, 2020 in Kerala. As the case count accelerated into March, the government put in place a multidimensional national lockdown beginning on March 22, which was dismantled in heterogeneous fashion across the country. The summer of 2020 was characterized by a range of adaptive policy regimes (Malani et al, 2020), with gradual reopenings and interspersed activity restrictions in response to changes in disease spread. By the end of 2020, seroprevalence surveys suggested cumulative infection rates in excess of 50% in many parts of the country.

A steady reopening into 2021 was followed by the worst wave of the pandemic thus far, triggering a new round of lockdowns around the country. At the time of writing, a third wave is feared and anticipated, and state governments are continuing to discuss the costs and benefits of restrictions on social behavior. A detailed analysis of the effects of such restrictions on mortalities caused by COVID-19 has again become crucial for current policy discussions^1^.

The effects of the various lockdown policies employed in India have not been systematically studied, in part because policies in 2020 were heterogeneous across space and not systematically documented. There has been wide debate on the matter, with some arguing that the national lockdown bought essential time for pandemic preparation, and others arguing that it accelerated the transmission of SARS-Cov-2 throughout the country by triggering the return journeys of millions of infected migrant workers. Most analyses of non-pharmaceutical interventions in India have focused on the national lockdown (See Narayanan and Saha (2021), Ceballos et al (2020) and Lee et al (2020)), or the green/yellow/red zone classification (See Poblete-Cazenave (2020), Ravindran and Shah (2020) and Beyer et al. (2020)) which immediately followed the end of the national lockdown, but was quickly abandoned by most states. Other studies have focused on the potential for specific NPIs, like information campaigns, to slow the spread of the virus (Banerjee et al, 2021).

We collected data on district-level non-pharmaceutical interventions (NPIs) in six Indian states with the highest infection counts as of the end of the summer of 2020, to assess the impact of NPIs on COVID transmission and fatality during the first half year of the pandemic in India.

## Data and Methods

Using media reports, we recorded the creation and removal of NPIs in the states of Andhra Pradesh, Bihar, Delhi, Karnataka, Maharashtra, and Tamil Nadu from March to August 2020^2^. This was one of the periods with the highest regional variability in NPIs, because states largely closed down all at once, but released their lockdowns in different dimensions in different places and times^3^. We recorded restrictions at the district-day level in 8 separate categories: border restrictions, curfews, industry closure, general lockdown, retail closures, school closures, temple closures, and transportation restrictions.

We coded each category of NPI in each district day, with a 1 indicating an NPI was in place and a zero otherwise. We then created an aggregate NPI index by taking the mean across all eight types of NPIs, effectively weighting each category of NPI equally. Maximal restriction was thus represented by 1, and minimal by 0.

We matched the district-level NPI data to district-level Google Mobility Trends, which is a coarse measure of individual travel behavior^4^, and to official counts of COVID positive tests and fatalities (aggregated by covid19india.org).

We ran linear regression models to estimate the relationship between NPIs and mobility trends (measured by change in percentage of population staying at home, from a weekday-specific baseline measured before the pandemic) at the district level with the following specification:

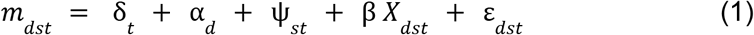

The outcome is *m*_*dst*_, the percentage change in population staying at home (from baseline levels) in district *d* and state *s* on date *t*. δ_*t*_, α_*d*_ and ψ_*st*_ are respectively date, district and state*date fixed effects. *X*_*dst*_ is the NPI index in district *d* in state *s* on date *t*. The date fixed effects control for country-wide trends in mobility over time as well as national policies (like the nationwide lockdown). The district fixed effects capture time-invariant characteristics of districts including level differences across districts in mobility trends and average NPI levels across the sample period. The state*date fixed effects control for state-level policy changes and aggregate trends in exposure to COVID-19, such as the mass return of migrants to Bihar following the national lockdown. The residual variation is thus driven by different policies in different districts of the same state, controlling for daily state-level changes in infections, mobility, and NPIs. The coefficient of interest is β, which describes the partial relationship between the NPI index and the share of the population staying at home.

To study the effect of NPIs on virus transmission and infection, we focused on COVID deaths instead of COVID case counts, because under-reporting of deaths is believed to be less severe than under-reporting of COVID cases. We ran the model as in Equation 1, with COVID deaths as the dependent variable. We lagged COVID deaths by 14 days to account for mean time between infection and death.

This model identifies the partial cross-district relationship between NPIs and COVID deaths, controlling for average state and time effects. A challenge to the causal interpretation of this model is that NPI policy may change in response to anticipated infections. This is most striking in Maharashtra, where the initial COVID surge predated and indeed was the cause of increasing NPIs in Maharashtra. The model is most plausibly interpreted causally in cases where NPIs were put in place in advance of the first wave of infections. Given that model interpretation may vary by state, we estimate the model on a state-by-state basis, in addition to the combined model above.

We also examined a second model, where we entered a separate binary variable for each category of NPI separately, to assess whether some categories of NPIs were more or less associated with infection spread.

According to our measure, NPI policies in Delhi were enacted at the state-level only; we therefore exclude Delhi from the district-level analysis.

## Results

Table 1 Column 1 shows the association between NPIs and the share of the population staying at home. The combined estimate suggests that the full set of NPI caused the share of time spent at home to increase by 8.8 percentage points relative to the baseline. In the state-level analysis, all states except Tamil Nadu show a large and statistically significant relationship between the NPI index and the population staying at home. Appendix Figure 1 shows the state-level time series of the average NPI index across districts against the stay-at-home measure. The high correlation between these measures is evident.

**Table 1:** Regression results with district and date fixed effects estimating the effect of NPI intensity. Dependent variables are stay-at-home rates in column 1 and death growth rates in column 2.

|  | Effect of NPI on share of time<br>spent at home<br>(1) | Effect of NPI on growth rate of<br>district-level number of deaths<br>(2) |
| --- | --- | --- |
| Bihar | 11.13565*** | 0.30705* |
|  | (0.91799) | (0.16114) |
| Tamil Nadu | -1.18415 | -0.77303*** |
|  | (0.88819) | (0.10871) |
| Karnataka | 17.24369*** | -0.96361*** |
|  | (2.12993) | (0.30235) |
| Andhra Pradesh | 3.91016** | -1.16492*** |
|  | (1.94849) | (0.34) |
| Maharashtra | 26.83596*** | 1.46366*** |
|  | (1.50302) | (0.17698) |
| Combined | 8.81115*** | -0.11809 |
|  | (0.5676) | (0.07558) |
| Combined (no MH) |  | -0.57628*** |
|  |  | (0.08243) |

Standard errors in parentheses
\* $p < 0.05$ , \*\* $p < 0.01$ , \*\*\* $p < 0.001$

This analysis establishes that NPIs (according to our original index) indeed had an impact on individual behavior. Note, however, that mobility is an imperfect measure of exposure to COVID risk. Actual risk is more likely associated with changes in household structure and exposure to crowds, which can take place even in the absence of substantial mobility. Similarly, individuals could reduce exposure to crowds substantially without changing their mobility by much. Therefore, we examine the effect of NPIs on COVID deaths directly rather than using mobility as an intermediate measure.

Figure 1 shows the pattern of NPIs plotted against the growth rate in COVID deaths in each of the six states. The NPI measure first moves in tandem across all states, rising from zero to one with the national lockdown on March 22. It then declines at different rates and times in different states (and districts, not shown).

In all states, the first surge in deaths is coincident with the national lockdown, reflecting that the lockdown was a policy response to the initial outbreak. In Maharashtra, where the initial outbreak was the most severe (with seroprelavence in excess of 50% in parts of Mumbai), this was the only major infection wave; in the other states, the majority of infections occurred after the initial lockdown ended. The graphs show that the major components of the lockdown were lifted at different times in different states; this is the primary variation that is exploited by our analysis below, since the initial rise in case growth and NPIs had similar timing in all states. In states other than Maharashtra the graphs show a broad pattern of accelerating deaths following release of the NPIs.

**Figure 2:**
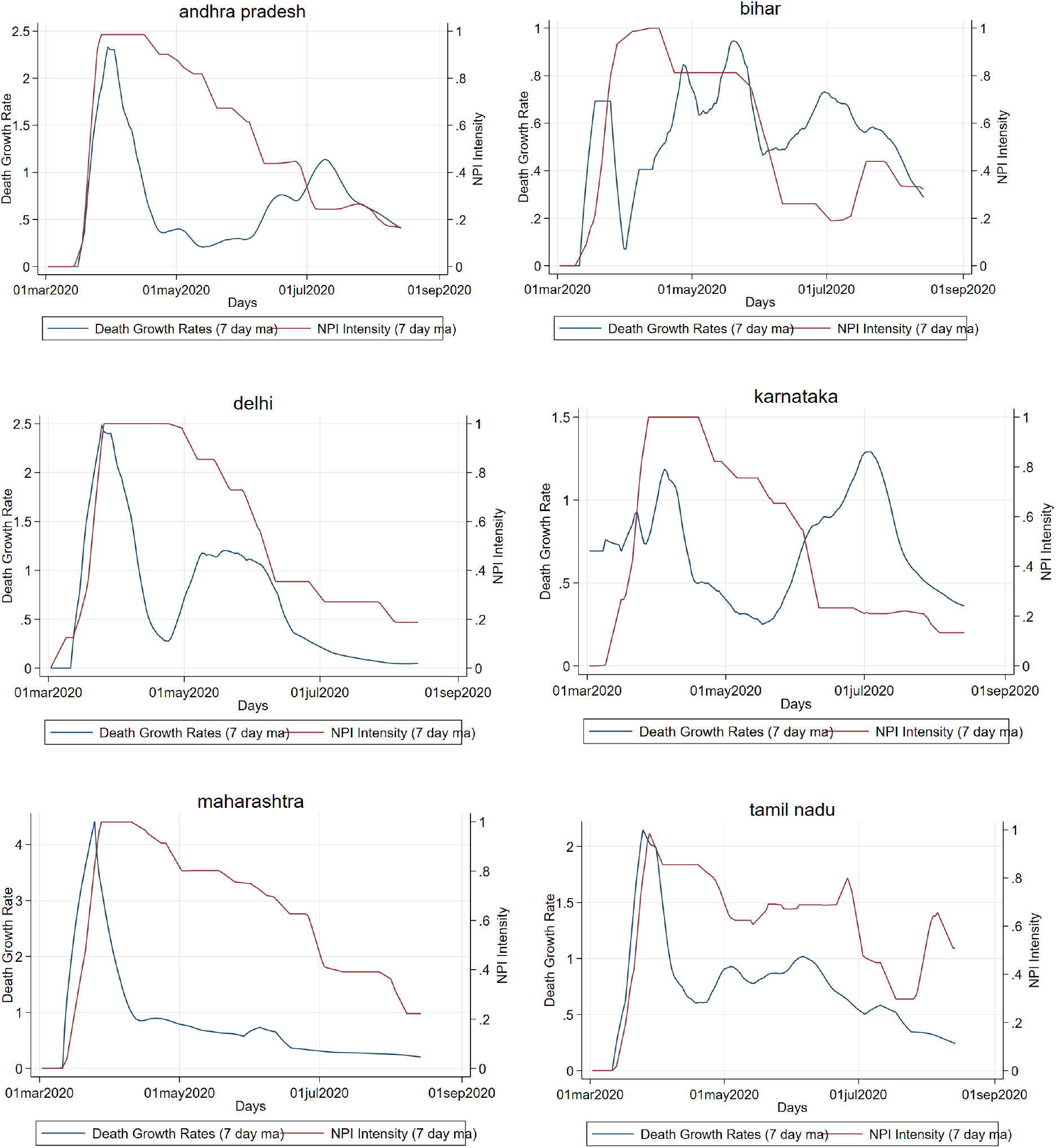
7 Day moving averages of death growth rates and NPI intensity in six states

Column 2 of Table 1 examines these results systematically in a regression framework. There is a large and highly statistically significant effect of NPI removal on growth in death rates in Andhra Pradesh, Karnataka, and Tamil Nadu. The coefficients on these states suggest that a complete removal of NPIs would increase the 14-day growth rate by 100 log points, or 65%.

In Maharashtra, the point estimate is positive and significant, reflecting the fact that the majority of deaths occurred coincidentally with the national lockdown. This is evidently a reverse causal relationship, and for this reason we remove Maharashtra from the later analysis below. In Bihar the estimate is small, positive, and marginally significant. It is possible that NPIs were ineffective in Bihar; an alternate explanation is that the coincident return of migrant workers spurred infection growth just as the first set of NPIs was being put in place (Burlig et al, 2021).

The combined estimate across six states is statistically insignificant, but becomes negative and strongly significant once Maharashtra is removed for the reasons above. The point estimate -0.577 implies that going from the full set of NPIs to none would cause a 44% reduction in the growth rate of deaths over the following 2-week period.

Table 2 disaggregates NPIs by category; we exclude Maharashtra because, as noted above, NPIs were put in place too late to have an effect on the major infection wave. The NPIs that are the most associated with subsequent reductions in mortality are curfews and closure of retail sectors and temples; industry closures, border closures, and transport restrictions are not associated with reductions in deaths. We exclude school closures, because these were closed from March 15 in all states through to the end of the sample, except in Maharashtra which is excluded from this analysis.^5^

**Table 2:**
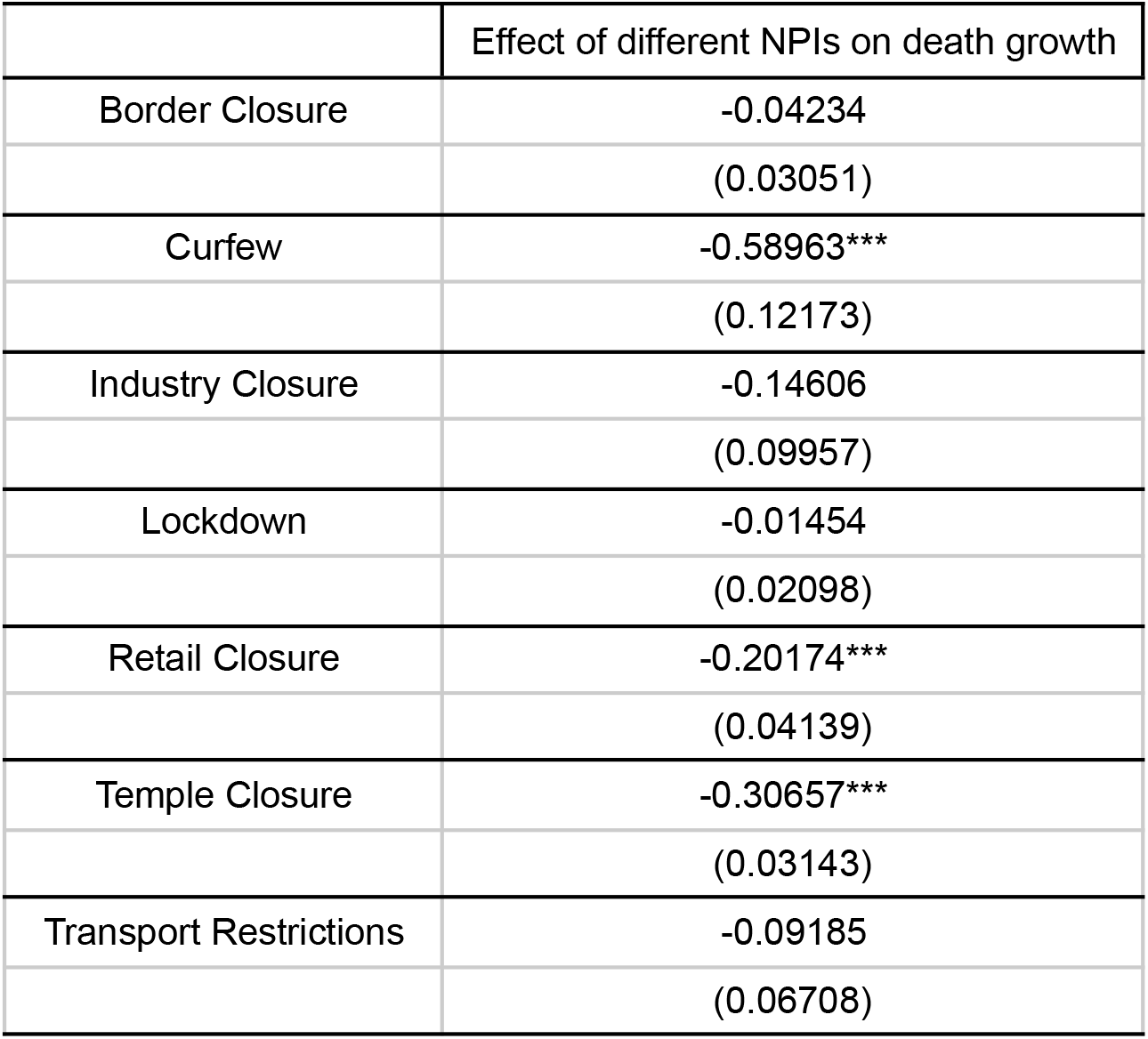
Regression results disaggregated by type of NPI, with district, date and state * date fixed effects

## Conclusion

Our results suggest that in 2020, regional NPIs were moderately effective at delaying COVID-19 infections in India. States and districts which re-opened more quickly after the pandemic lockdown faced earlier waves of the pandemic. Sustained curfews, retail and temple closures were particularly associated with delayed pandemic waves, perhaps because they were the key measures that prevented large gatherings and super-spreader events. Adaptive control in India may have been effective in flattening the pandemic curve and buying time for clinics to prepare for the pandemic, and there was considerable heterogeneity across states on this count.

Our study faces the same limitations as other analyses of NPIs. NPIs were not put in place randomly and thus their variation could reflect (incorrect) anticipation of a decrease in the intensity of the pandemic in some states. India’s data on COVID deaths is almost surely biased downward, though the evidence so far suggests the cross-state patterns in deaths have reflected actual differences in pandemic intensity.

Since various categories of NPIs also tend to be correlated, our disaggregated results on the impact of various NPI categories also need to be read with a grain of salt. Last, our approach was focused on regional NPI policies and we did not estimate spillovers of national policies through induced migration. Our open data on the categorized NPIs can facilitate future research on NPI effectiveness as better data on pandemic spread becomes available.

Our analysis predates India’s second COVID wave, which had daily death and case counts that were three times higher than the first wave that is studied here. It is of note, and consistent with our findings above, that the second wave was preceded by abandonment of most NPIs in many parts of the country, and a return to mass activities.

## Data Availability

All data were obtained from public sources. The complete replication code and underlying datasets can be found at github.com/devdatalab/paper-kalra-novosad-india-npi.

https://github.com/devdatalab/paper-kalra-novosad-india-npi

## Appendix

**Appendix Figure 1:**
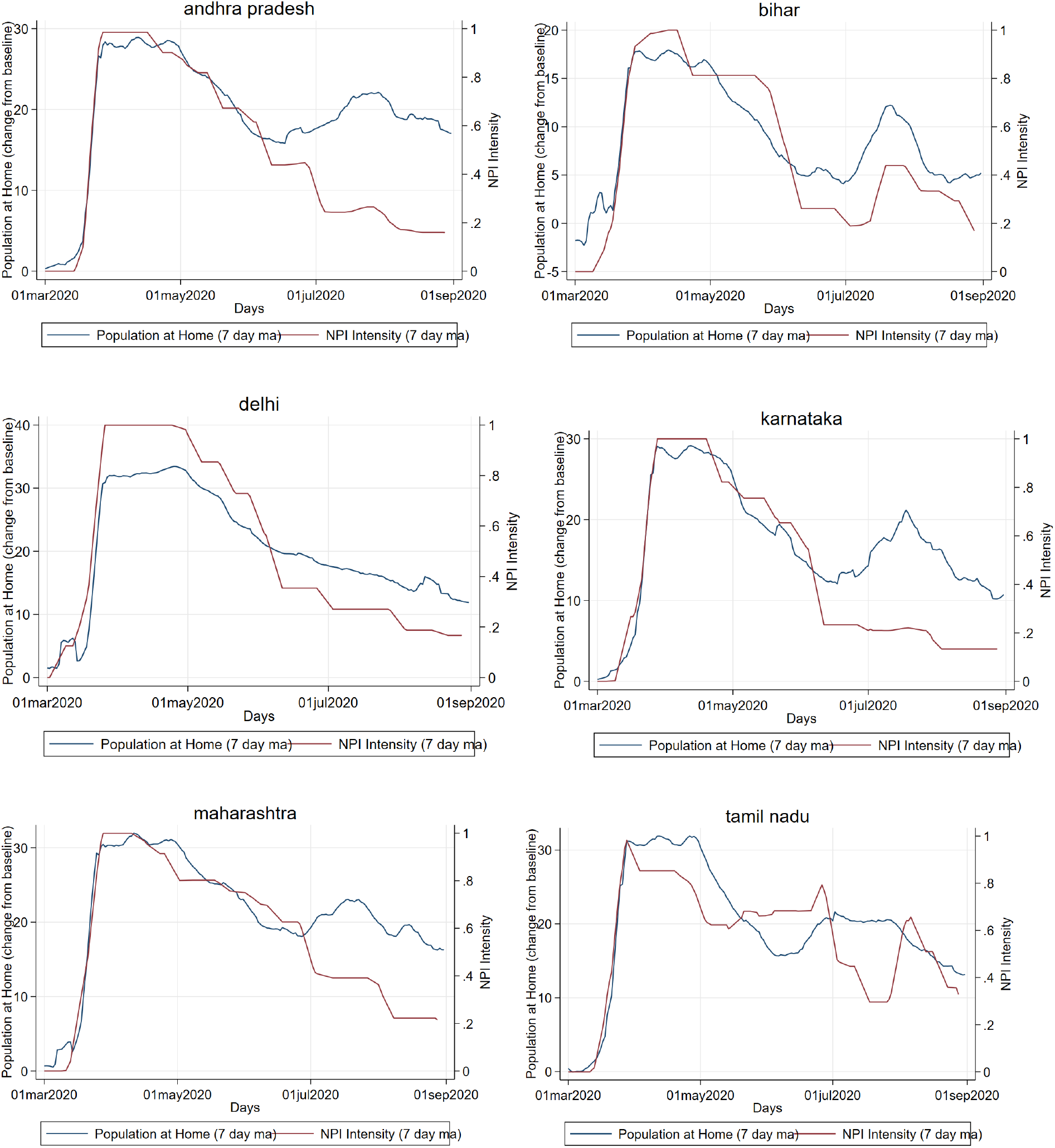
7 Day moving averages of percentage change in population staying at home and NPI intensity in six states

## Footnotes

1 We reviewed methods employed to conduct similar analysis across various contexts. Barro (2020) studies the relationship between non-pharmaceutical interventions (NPI) and mortality in the US during the 1918-19 Influenza Pandemic. Flaxman et al (2020) estimates the effects of NPIs on COVID-19 in Europe.

2 Hsiang et al (2020) also follow similar methods to collect NPI data in six countries (not including India), using Wikipedia, government sources, and news articles.

3 Gupta et al (2021) measure intensity of government restrictions using the Oxford COVID-19 Government Response Tracker. However, this dataset provides measures of severity at the All-India level only, and not at the sub-national levels, and is thus not informative about local policies, which formed the bulk of the post-lockdown response.

4 We use district-level google mobility trends because mortality data is also aggregated at the district level all over India. Sheng et al (2021) perform more granular analysis within Mumbai using smart phone location data from Infinite Analytics, a marketing analysis company.

5 The absence of effect on lockdowns is difficult to interpret since lockdowns effectively comprise a combination of the above measures; one interpretation is that lockdowns had limited effect over and above curfews, retail and temple closures. In other words, those three components may have been the most effective aspects of the lockdowns. However, limited data, policy endogeneity and collinearity all limit our certainty.

## Notes

### Competing Interest Statement

The authors have declared no competing interest.

### Funding Statement

No external funding was received for this project.

### Author Declarations

This project uses publically available data sources.

## References

Banerjee, Abhijit, Marcella Alsan, Emily Breza, Arun G. Chandrasekhar, Abhijit Chowdhury, Esther Duflo, Paul Goldsmith-Pinkham, and Benjamin A. Olken. Messages on COVID-19 prevention in India increased symptoms reporting and adherence to preventive behaviors among 25 million recipients with similar effects on non-recipient members of their communities. No. w27496. National Bureau of Economic Research, 2020.

Barro, Robert J. Non-pharmaceutical interventions and mortality in US cities during the great Influenza pandemic, 1918–1919. No. w27049. national bureau of economic Research, 2020.

Beyer, Robert, Tarun Jain, and Sonalika Sinha. “ Lights out? covid-19 containment policies and economic activity.” COVID-19 Containment Policies and Economic Activity, 2021.

Burlig, Fiona, Anant Sudarshan, and Garrison Schlauch. The impact of domestic travel bans on COVID-19 is nonlinear in their duration. No. w28699. National Bureau of Economic Research, 2021.

Ceballos, Francisco, Samyuktha Kannan, and Berber Kramer. “ Impacts of a national lockdown on smallholder farmers’ income and food security: Empirical evidence from two states in India.” World Development 136 (2020): 105069.

Flaxman, Seth, Swapnil Mishra Axel Gandy, H. Juliette T. Unwin, Thomas A. Mellan, Helen Coupland, Charles Whittaker et al. “ Estimating the effects of non-pharmaceutical interventions on COVID-19 in Europe.” Nature 584, no. 7820 (2020): 257–261.

Gupta, Arpit, Anup Malani, and Bartek Woda. Explaining the Income and Consumption Effects of COVID in India. No. w28935. National Bureau of Economic Research, 2021.

Hsiang, Solomon, Daniel Allen, Sébastien Annan-Phan, Kendon Bell, Ian Bolliger, Trinetta Chong, Hannah Druckenmiller et al. “ The effect of large-scale anti-contagion policies on the COVID-19 pandemic.” Nature 584, no. 7820 (2020): 262–267.

Lee, Kenneth, Harshil Sahai, Patrick Baylis, and Michael Greenstone. “ Job loss and behavioral change: the unprecedented effects of the India lockdown in Delhi.” University of Chicago, Becker Friedman Institute for economics working paper 2020–65 (2020).

Malani, Anup, Satej Soman, Sam Asher, Paul Novosad, Clement Imbert, Vaidehi Tandel, Anish Agarwal et al. Adaptive control of COVID-19 outbreaks in India: local, gradual, and trigger-based exit paths from lockdown. No. w27532. National Bureau of Economic Research, 2020.

Narayanan, Sudha, and Shree Saha. “ Urban food markets and the COVID-19 lockdown in India.” Global Food Security 29 (2021): 100515.

Poblete-Cazenave, Ruben. “ The great lockdown and criminal activity–evidence from Bihar, India.” 2020.

Ravindran, Saravana, and Manisha Shah. Unintended consequences of lockdowns: covid-19 and the shadow pandemic. No. w27562. National Bureau of Economic Research, 2020.

Sheng, Jaymee, Anup Malani, Ashish Goel, and Purushotham Botla. Does Mobility Explain Why Slums Were Hit Harder by COVID-19 in Mumbai, India?. No. w28541. National Bureau of Economic Research, 2021.

